# Effectiveness of Pfizer-BioNTech Vaccine Against COVID-19 Associated Hospitalizations among Lebanese Adults ≥75 years- Lebanon, April-May 2021

**DOI:** 10.1101/2022.01.19.22269514

**Authors:** Zeina Farah, Nadine Haddad, Hala Abou El Naja, Majd Saleh, Pamela Mrad, Nada Ghosn

## Abstract

**Introduction:** In Lebanon, the nationwide vaccination against COVID-19 was launched in February-2021 using Pfizer-BioNTech vaccine and prioritizing elderly, persons with comorbidities and healthcare workers. Our study aims to estimate the post-introduction vaccine effectiveness (VE) of Pfizer-BioNTech in preventing COVID-19 hospitalization among ≥75 years in Lebanon.

**Methods:** A case-control study design was used. Cases were Lebanese, ≥75 years and hospitalized with positive PCR result during April-May 2021. Cases were randomly selected from the COVID-19 database of the Epidemiological Surveillance Unit(ESU) at the Ministry of Public Health(MOPH). Each case was matched by age and locality to 2 controls. Controls were hospitalized, non-COVID-19 patients with negative PCR result, randomly selected from the MOPH hospital admission database. VE was calculated for fully and partially vaccinated, using multivariate conditional logistic regression analyses.

**Results:** 345 cases and 814 controls were recruited. Half were females, with a mean age of 83 years. 14 cases(5%) and 143 controls(22%) were fully vaccinated. Bivariate analysis showed significant association with: gender, month of confirmation/admission, general health, chronic medical conditions, main income source and living arrangement. After adjusting for month of admission and gender, multivariate analysis yielded a VE of 82% (95%CI = 69%-90%) against COVID-19 associated hospitalization for those fully vaccinated and 53% (95%CI = 23%-71%) for those partially vaccinated (≥14 days of first or within 14 days of second dose).

**Conclusions:** Our study showed that Pfizer-BioNTech vaccine is effective in reducing risk for COVID-19– associated hospitalization in Lebanese elderly(≥75 years). Additional studies are warranted to explore vaccine effectiveness in reducing hospitalization in younger age groups, as well as in reducing covid-19 infections.

## Introduction

Since its emergence in December 2019, the severe acute respiratory syndrome coronavirus 2 (SARS-CoV-2) has taken a tremendous toll on the population worldwide. By 4 May 2021, there have been over 153 million cases and 3.2 million deaths globally from COVID-19(1). The implemented non-pharmacological interventions affected the daily lives of billions around the world, resulting in devastating socio-economic repercussions - the biggest challenge that humanity ever faced since World War II(2).

In response to this pandemic, considerable efforts were put globally into developing effective and safe drugs and vaccines against SARS-CoV-2(3). Candidate vaccines were introduced with differing indications, contra-indications and adverse events - each with a specific efficacy in preventing SARS-CoV-2 infection, severe outcomes, and death(4). Hence, it is of a paramount importance to evaluate the post-introduction effectiveness of the approved and marketed vaccines.

In Lebanon, a total of 529,205 COVID-19 confirmed cases and 7,368 related deaths were reported as of 4 May 2021 since the detection of the first case on 21 February 2020(5). In January 2021, the Lebanese Ministry of Public Health(MOPH) issued its National Deployment and Vaccination Plan for COVID-19 vaccines(NDVP) in which priority target groups were selected based on specific risk factors: age, comorbidity, and occupation in line with the World Health Organization(WHO) Strategic Advisory Group of Experts on Immunization (SAGE) recommendations(6). On 14 February 2021, almost a year after the beginning of the outbreak, the national vaccination was rolled out upon the arrival of the first batch of Pfizer-BioNTech vaccine(7). On 24 March 2021, the country received the first batch of AstraZeneca vaccine, hence putting forward the deployment of the national vaccination plan. As of 6 May 2021, a total of 498,722 vaccine doses of various types (Pfizer-BioNTech, AstraZeneca, Sputnik V, SinoPharm) were administered as reported through the Inter-Ministerial and Municipal Platform for Assessment, Coordination and Tracking (IMPACT) platform(7). Pfizer-BioNTech vaccine was used for residents aged 75 and above with a time interval of 21 days between the 2 doses and a coverage of 37% for 2 doses, as of 6 May 2021.

As the nationwide vaccination progresses, it is vital to study the vaccine effectiveness in the community as emphasized in the NDVP. Particularly, the national plan mentions the role of the Epidemiological Surveillance Unit(ESU) in studying vaccine effectiveness in order to guide vaccination policies and Public Health and Social Measures(6).

Hence, our study aimed to estimate the post-introduction effectiveness of Pfizer-BioNTech COVID-19 vaccine against COVID-19 hospitalization among Lebanese adults ≥75 years.

## Methods

### Study design

A case-control (CC) study design was conducted using structured questionnaires conducted via phone call interviews(10). Our sample size was calculated using the precision method with 90% vaccine effectiveness for full vaccination and 40% vaccine coverage, a ±5% precision and a type 1 error of 0.05. A minimum sample size of 318 cases and 636 controls was needed(8). The minimal sample size was multiplied by 50% to account for non-response and refusals. Proportionate matching with 1:2 cases to control ratio was conducted according to age group ([75-85[; [85-95)) and locality (governorate) of residence.

### Study population

The COVID-19 surveillance database of the ESU-MOPH was screened to select confirmed cases reported as Lebanese, ≥75 years, hospitalized and diagnosed with COVID-19 by Real Time-Polymerase Chain Reaction between April and May 2021. A random sample of 742 cases was drawn from the selected sampling frame and contacted to verify whether they meet our study inclusion criteria. Cases not meeting one of the inclusion criteria were discarded from our analysis.

Controls were selected randomly from the MOPH hospital admission database(of uninsured individuals covered by the MOPH) which comprises around 50% of all hospitalizations in Lebanon(9). The selected controls were Lebanese, ≥ 75 years old and hospitalized between April and May 2021 with admission diagnosis related to all International Classification of Diseases (ICD-10) chapters excluding the COVID19 code. During our study period, any hospitalization due to causes other than COVID-19 required a negative RT-PCR prior to admission, so it was assumed that the negative status of controls is ascertained. Controls were later excluded if the investigation showed they do not meet one of our study inclusion criteria.

### Variables

A structured questionnaire was used including socio-demographic information(age, gender, place of residence and main source of income), living conditions(number of household members, number of rooms and living arrangement), health conditions in the 12-month period prior to admission(perception of general health status, presence of comorbidities and ability to walk and climb), hospitalization status(duration of hospitalization, admission to Intensive Care Unit(ICU), duration of stay at ICU, oxygen therapy, intubation, and discharge status) in addition to cognitive variables (ability to read and performing calculations). Further, crowding index was computed by dividing the number of household members by the number of rooms. The date of PCR test result confirmation was available for all cases. However, the date of admission was used for controls as it was available in the MOPH admission database. This time variable was considered a confounding factor as both the vaccination coverage and COVID-19 incidence show time trends during the study period(7,10). Three categories for hospital stay were generated: <3 days, 3 to 7 days and above 7 days.

Participants self-reported their vaccination status by indicating their vaccination dates according to the received SMS from the MOPH vaccination platform. Vaccination data was considered only for participants reporting exact dates of vaccination. Vaccination status included 4 categories: 1)”Unvaccinated” defined as no receipt of Pfizer-BioNTech vaccine before confirmation/admission, 2)Single-dose vaccinated <14 days before confirmation/admission, 3)”Partially vaccinated” defined as receipt of 1 dose of Pfizer-BioNTech vaccine ≥14 days before confirmation/admission or 2 doses, with the second dose received <14 days before confirmation/admission 4)”Fully vaccinated” defined as receipt of 2 doses of the vaccine with the second dose received ≥14 days before confirmation/admission.

### Data management

Collected data was digitalized using DHIS2 tracker program and analyzed using R version 4.0.4 and R studio version 1.4.1103. For the VE analysis, fully and partially vaccinated participants were compared to unvaccinated subjects. Univariate descriptive statistics was used to assess the distribution of covariates among participants and identify potential confounding factors. Characteristics of cases and controls were compared using Chi-square tests or Fisher’s exact tests for categorical variables and Student’s t-test or Wilcoxon rank-sum tests for continuous variables. For the final selection of potential confounders to include in the logistic regression model, the “change-in-estimate” approach was used (8). Covariates whose adjustment changed the crude odds ratio by ≥5% were included in the final models. VE was estimated using conditional logistic regression following the below formula(8):

VE= (1 -matched, adjusted odds ratio for vaccination) x 100%

The 95% Confidence Intervals(CIs) of VE were calculated as 1 - CI_OR_, where CI_OR_ is the confidence interval of the odds ratio estimates.

### Ethical approval

The study was approved by the Institutional Review board of the Rafik Hariri University Hospital. Informed consent was obtained from study subjects prior to participation.

## Results

### Study population

Between April 1^st^ and May 31^st^ 2021, 742 cases and 1,561 controls were contacted for the study. However, due to either being excluded for not fitting case/control definition(34%; 23%), no reply(16%; 20%) or refusal to participate(4%; 5%), the retained number of participants was 345 cases to 814 matched controls(Figure 1).

**Figure 1:**
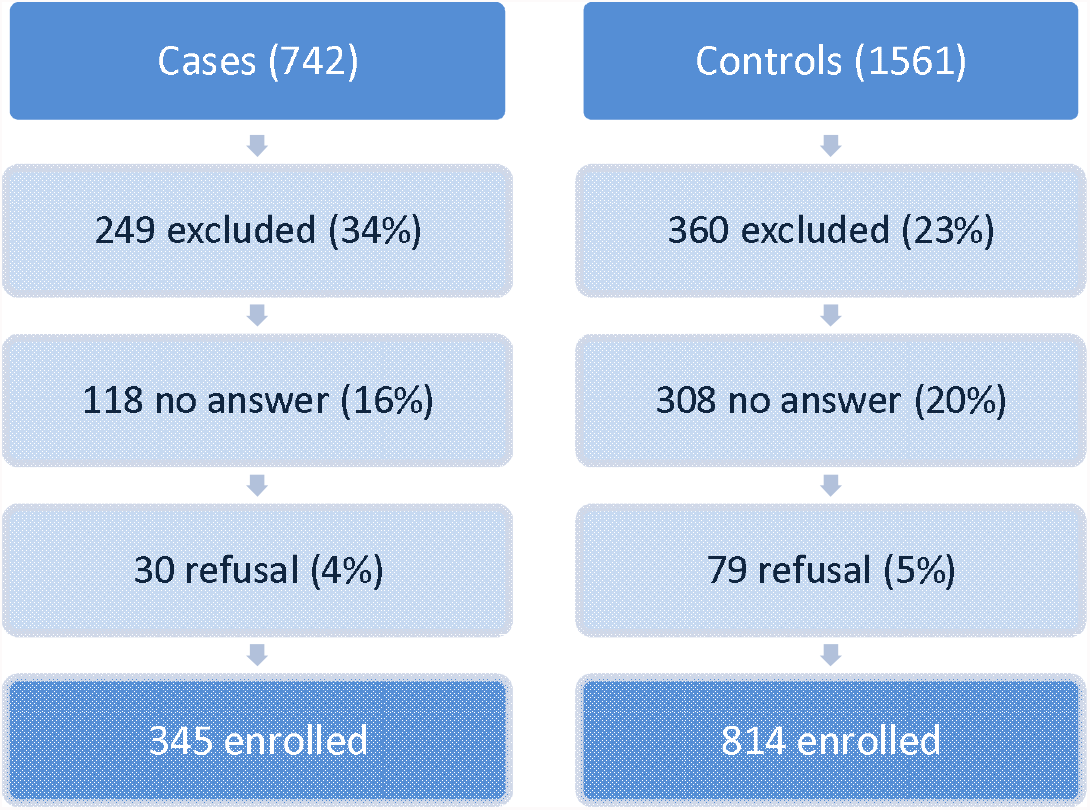
Selection of study participants and sample size, VE CC study, ≥75 years, Apr-May2021, Lebanon

The mean age of participating cases and controls was 83.1±5.6 and 82.8 ±5.7 years, respectively. The highest proportion of cases and controls resided in Mount-Lebanon governorate (37%; 39%). Cases had an equal proportion of females and males (50%); however, the control group had a significantly higher proportion of females (57%) (*P=*.03) (Table 1).

**Table 1:**
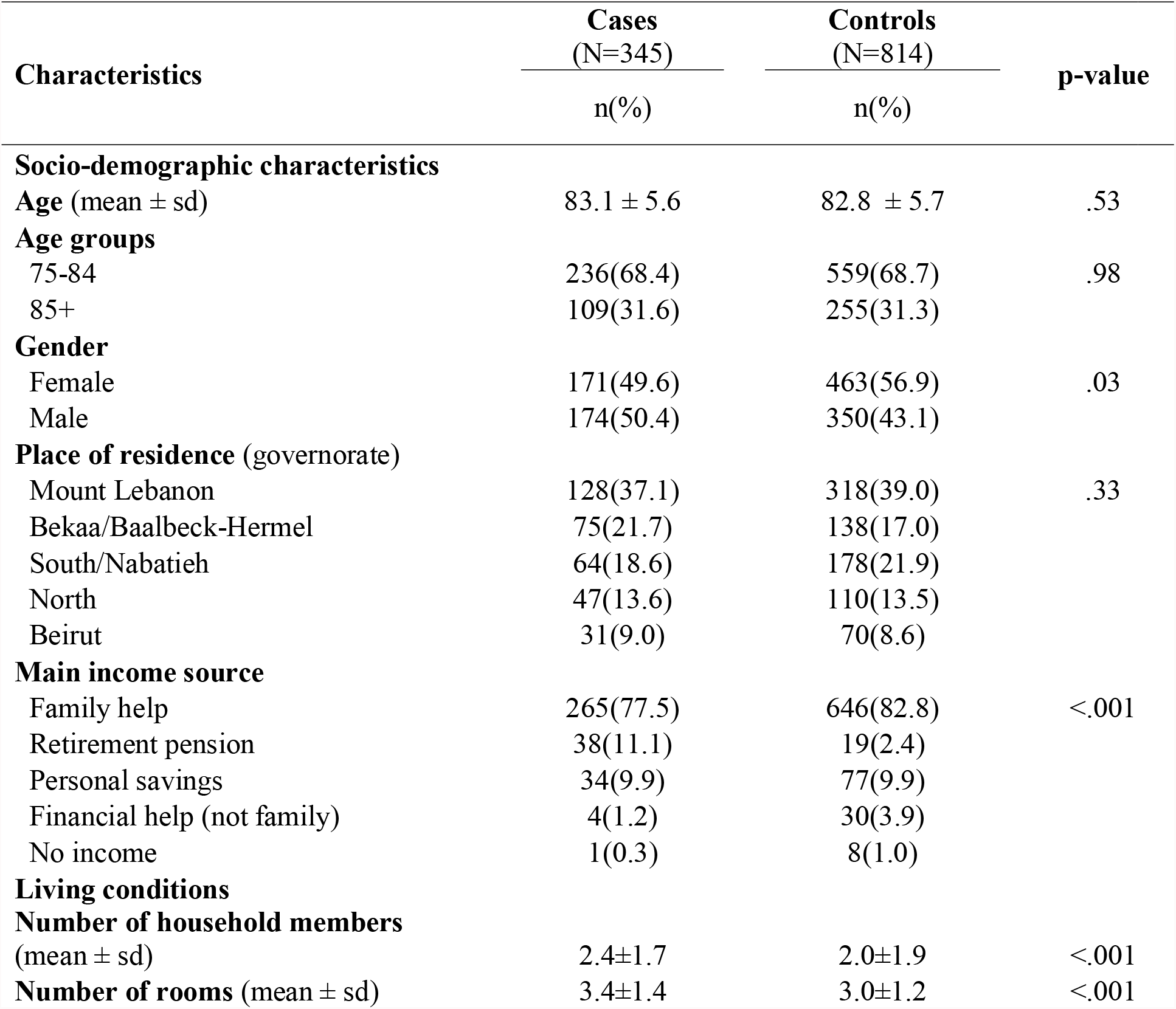
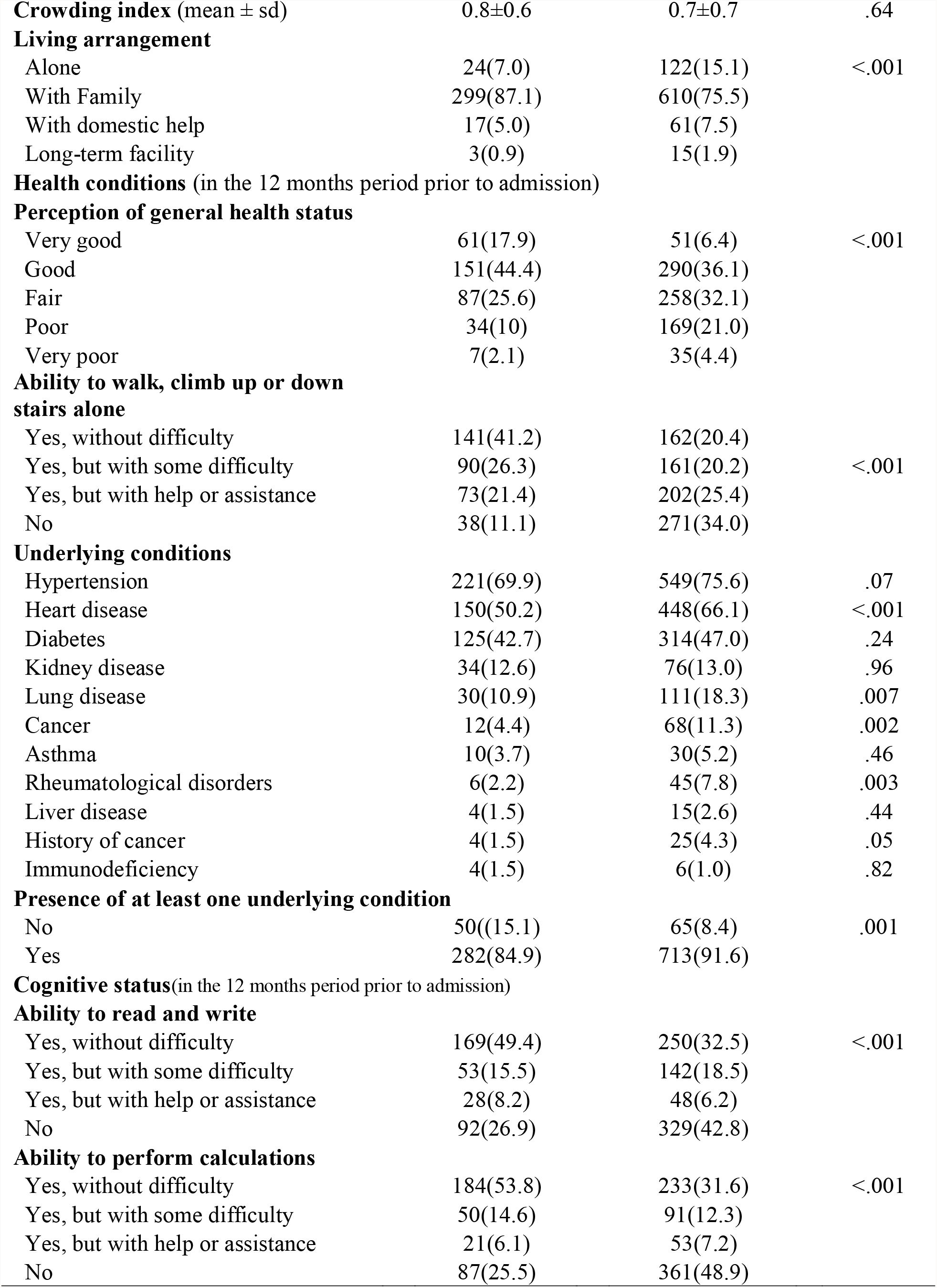
Characteristics of Lebanese hospitalized COVID-19 case-patients and controls aged ≥75 years, April-May 2021

Concerning household arrangement, cases had a greater number of rooms (3.4±1.4) compared to controls (3.0±1.2) and a higher number of household members (2.4±1.7) compared to controls (2.0±1.9) (*P* <.001). Most cases and controls (87%; 76%) lived with family members. No significant differences were noted for the crowding index between our comparison groups (*P*=.64) (Table 1).

For the vast majority in both groups, family support was the main source of income. However, cases (11%) had more retirement pensions than controls (2%) (Table 1).

Cases (54%, n= 184) were significantly more likely than controls (32%, n=233) to perform calculations without difficulty. They were also more likely to read and write without difficulty (49% of cases as compared to 33% among controls, *P*<.001). On the other hand, the proportion of subjects reporting inability to read and write was higher among controls (43%, n=329) than cases (27%, n=92) (Table 1).

The general health status of the cases and controls in the 12 months prior to admission was also significantly different (*P*<.001) and was mostly good (44% for cases and 36% for controls) followed by fair health status (26% cases and 32% for controls). 41% of cases reported no difficulty walking, climbing up and down stairs prior to admission, while 34% of controls did not have the ability to conduct this task (Table 1).

Majority of cases and controls had at least one underlying condition (85%; 92%) (*P*=.001). The underlying conditions reported for both cases and controls were mostly hypertension (70%; 76%) followed by heart diseases (50%;66%), and diabetes (43%; 47%) respectively (Table 1).

### Hospitalization

As for hospitalization information, the mean duration of hospital stay was significantly higher among cases (11.1±9.3 days) compared to controls (6.0±7.0 days). In particular, the length of stay was mostly >7 days for cases (53%) while 3-7 days for controls (60%). Majority of cases required ICU admission (67%) with a longer duration of stay (8.5±7.9 days) while 25% of controls were admitted to ICU (*P*<.001) (Table 2).

**Table 2:** Hospitalization data of Lebanese hospitalized COVID-19 case-patients and controls aged ≥75 years, April-May 2021

| Characteristics | Cases<br>(N=345) | Controls<br>(N=814) | p-value |
| --- | --- | --- | --- |
|  | n(%) | n(%) |  |
| **Hospitalization status** |  |  |  |
| **Month of confirmation/**<br>**admission \*** |  | - |  |
| April | 271(78.6) | 464(57.0) | <.001 |
| May | 74(21.4) | 350(43.0) |  |
| **Duration of**<br>**hospitalization (days)** | 11.1±9.3 | 6.0±7.0 | <.001 |
| **Hospital stay** |  |  |  |
| <3 days | 31(9.6) | 160(22.3) | <.001 |
| 3-7 days | 120(37.0) | 426(59.5) |  |
| >7 days | 173(53.4) | 130(18.2) |  |
| **Admission to ICU** |  |  |  |
| Yes | 224(66.9) | 180(24.6) | <.001 |
| No | 111(33.1) | 551(75.4) |  |
| **Duration of stay at ICU** | 8.5±7.9 | 5.8±5.8 | <.001 |
| **Oxygenotherapy** |  |  |  |
| Yes | 275(90.2) | 175(25.8) | <.001 |
| No | 30(9.8) | 503(74.2) |  |
| **Mechanical ventilation** |  |  |  |
| Yes | 107(46.3) | 40(6.1) | <.001 |
| No | 124(53.7) | 611(93.9) |  |
| **Discharge status** |  |  |  |
| Alive | 153(48.3) | 711(89.8) | <.001 |
| Death | 164(51.7) | 81(10.2) |  |
| **Cause of death<sup>§</sup>** |  |  |  |
| Due to COVID-19 | 136(93.4) | 0(0) | <.001 |
| Due to other causes | 9(6.2) | 70(100) |  |

\* Month of confirmation for cases and admission for controls
<sup>§</sup> Among deaths

Furthermore, cases were significantly more likely than controls to require oxygen therapy (90%) and intubation (46%) (*P*<0.001). Death upon discharge was significantly higher among cases (52%) compared to controls (10%) (*P*<0.001). Among cases, the majority were due to COVID-19(93%) (Table 2). Of note, no change in the above results was found when restricting the Univariate analysis to participants with complete vaccination data.

### Vaccination effectiveness

Majority of cases and controls reported zero dose of vaccination (81%; 63%), 13% of cases reported having received one dose of vaccine compared to 11% for controls, and 5% of cases reported having two doses compared to 27% of controls. Majority of cases and controls reported no side effects following 1^st^ dose (66%; 82%) and second dose (67%; 81%) respectively (*P=*.002 and .13 respectively) (Table 3).

**Table 3:**
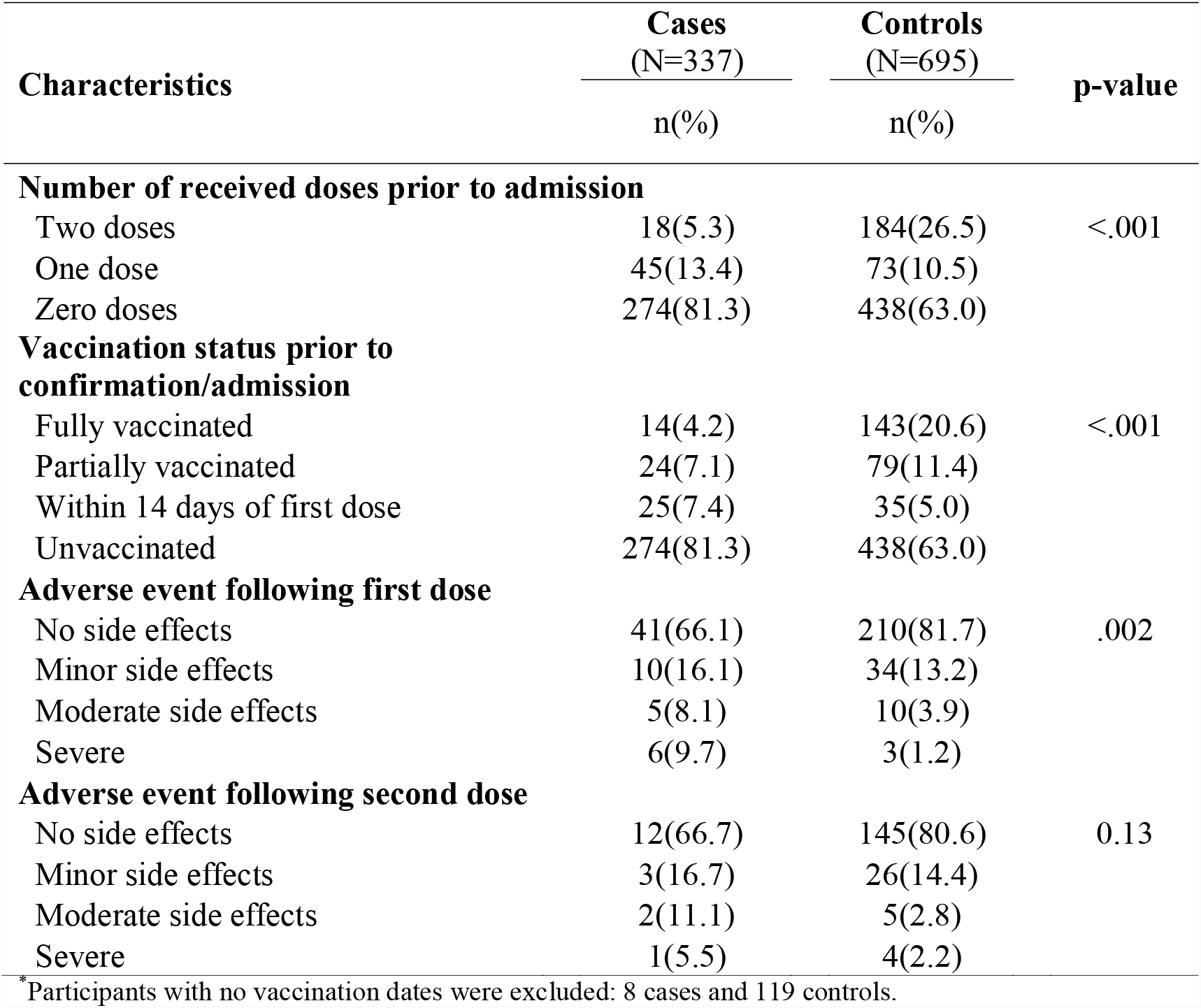
Vaccination data of Lebanese hospitalized COVID-19 case-patients and controls aged ≥75 years, April-May 2021^*^

| Characteristics | Cases<br>(N=337) | Controls<br>(N=695) | p-value |
| --- | --- | --- | --- |
|  | n(%) | n(%) |  |
| **Number of received doses prior to admission** |  |  |  |
| Two doses | 18(5.3) | 184(26.5) | <.001 |
| One dose | 45(13.4) | 73(10.5) |  |
| Zero doses | 274(81.3) | 438(63.0) |  |
| **Vaccination status prior to confirmation/admission** |  |  |  |
| Fully vaccinated | 14(4.2) | 143(20.6) | <.001 |
| Partially vaccinated | 24(7.1) | 79(11.4) |  |
| Within 14 days of first dose | 25(7.4) | 35(5.0) |  |
| Unvaccinated | 274(81.3) | 438(63.0) |  |
| **Adverse event following first dose** |  |  |  |
| No side effects | 41(66.1) | 210(81.7) | .002 |
| Minor side effects | 10(16.1) | 34(13.2) |  |
| Moderate side effects | 5(8.1) | 10(3.9) |  |
| Severe | 6(9.7) | 3(1.2) |  |
| **Adverse event following second dose** |  |  |  |
| No side effects | 12(66.7) | 145(80.6) | 0.13 |
| Minor side effects | 3(16.7) | 26(14.4) |  |
| Moderate side effects | 2(11.1) | 5(2.8) |  |
| Severe | 1(5.5) | 4(2.2) |  |

\*Participants with no vaccination dates were excluded: 8 cases and 119 controls.

Taking fully vaccinated individuals as 14 days after second dose, 14 cases (4%) and 143 controls (21%) were fully vaccinated and the crude OR was 0.16 (95%CI = 0.09-0.28). After adjusting for month of admission and gender, multivariate analysis yielded an adjusted OR of 0.18 and a VE of 82% (95%CI = 69%-90%) (Table 3). For those partially vaccinated, the crude OR was 0.49 (95%CI = 0.30-0.79). After adjusting for month of admission, the adjusted OR was 0.47 and the vaccine effectiveness was 53% (95%CI = 23%-71%) (Table 4).

**Table 4:** Pfizer-BioNTech vaccine effectiveness^*^ against COVID-19 among Lebanese hospitalized COVID-19 case-patients and controls aged ≥75 years, April-May 2021 (n=1,032)

| Vaccination status | Cases | Controls | Unadjusted OR (95% CI) | VE against COVID-19 hospitalization (95% CI) |
| --- | --- | --- | --- | --- |
| **Fully vaccinated** | 14(4.2) | 143(20.6) | 0.16(0.09-0.28) | 82(69-90)* |
| **Partially vaccinated** | 24(7.1) | 79(11.4) | 0.49(0.30-0.79) | 53(23-71)** |

\*Adjusted for month of confirmation/admission and gender
\*\*Adjusted for month of confirmation/admission

On the other hand, there was no significant effect for receiving the first dose of the vaccine within 14 days of confirmation/admission (adjusted OR = 1.09 and 95% CI = 0.63-1.87).

## Discussion

Clinical trials have assessed the efficacy of Pfizer-BioNTech vaccine against COVID-19 associated infection; however, monitoring efficacy against COVID-19 associated hospitalization was not possible because hospitalization is a rare outcome among COVID-19 patients(11). Our study supports assessment of COVID-19 hospitalization among a high-risk group. In this analysis of Lebanese ≥ 75 years old hospitalized between April and May 2021, vaccination with Pfizer-BioNTech was significantly less likely among hospitalized patients with COVID-19 than other conditions. These findings are consistent with available evidence showing reduction in COVID-19 associated hospitalization among vaccinated patients compared to unvaccinated subjects (12)

In this study, multivariate analysis of hospitalized patients ≥ 75 years revealed that Pfizer-BioNTech vaccine was associated with significant protection against COVID-19 hospitalization. Effectiveness was 82% among elderly 75 years and above who were fully vaccinated (14 days after second dose) and 53% among those who were partially vaccinated (having received 1 dose of vaccine ≥14 days before confirmation/admission or 2 doses, with the second dose received <14 days after confirmation/admission).

Our findings suggest that two doses of Pfizer-BioNTech vaccine at least 14 days after vaccine administration provided a substantial level of protection (82% VE) against hospitalization for elderly individuals (≥75years) in Lebanon between April and May 2021. These results are consistent with previous studies showing similar results, mainly two studies conducted in the United States(US) during the period March-July and February-August 2021 targeting adults, showing the VE of mRNA vaccines (Pfizer-BioNTech and Moderna) against hospitalization to be 86% (95%CI = 82%-88%) and 80% (95% CI = 68%-87%) among fully vaccinated aged 65 years and above (13,14).

Moreover, in assessing the impact of one dose of the vaccine, no significant effectiveness within the 14 days of the first dose was detected. This is also in line with results from other studies showing no significant effect in the 14 days after the first dose (11,15).

However, our findings are lower than the VE reported in some studies assessing the effectiveness of mRNA vaccines against COVID19 hospitalization mainly two studies conducted during January-March and March-August 2021. These studies targeted adults in the US showing the VE of mRNA vaccines (Pfizer-BioNTech and Moderna vaccines) for fully vaccination to be 94% (95% CI = 49%–99%) for adults ≥65years and the VE of Pfizer-BioNTech vaccine to be 91%(95% CI = 88%-93%) among adults ≥18years, respectively (11,16).

As for the effectiveness of partial vaccination, our findings were lower than those reported in a study conducted in the US showing the Pfizer-BioNTech vaccine VE to be 64% (95%CI=28%-82%) against hospitalization (after 14 days of first dose or within 14 days of second dose) among adults ≥ 65 years(11). It was also lower than those reported in other studies assessing the VE against hospitalization after 14 days of one Pfizer-BioNTech dose, showing a VE of 71% (95%CI=47%-91%) among elderly ≥80 years in the UK and a VE of 70% (95%CI=60%-77%) among people ≥16 years in Canada (17,18)

The difference between the study results could be due mostly to the mean age group of this study which is around 83 years, higher in comparison to the other studies. Moreover, the fact that some of these studies were assessing a combination of mRNA vaccines, not only the Pfizer-BioNTech vaccine could have affected the comparison. Other factors to mention include the difference in the study design, evaluated population and inclusion criteria between the different studies as well as the variability in unmeasured confounding factors.

Moreover, our study took place between April and May 2021 where the dominant variant in Lebanon is suggested to be Alpha. In Lebanon, genomic sequencing began in June with Alpha variant being the dominant circulating virus (19); SARS-CoV2 delta variant took over and became the dominant circulating virus in Lebanon starting July 2021. The fact that some VE studies were targeting delta variant, this might have led to the observed differences in VE values when comparing our results to other studies. Therefore, it is very crucial to interpret VE results and cautiously compare them with other international studies taking into account SARS-CoV2 circulating variants at time of study period.

Furthermore, in our study design, targeting elderly individuals above 75 years for both cases and controls, might have affected vaccine intake as well as exposure to infection in the sense that decreased mobility of elderly individuals might have affected the accessibility to vaccination; likewise, elderly individuals, due to their vulnerable conditions, might have limited their exposure to their bubble, hence decreasing the risk of exposure to infection during the pandemic. Any of these directions might have biased our VE estimates.

Additionally, majority of cases and controls reported having zero dose of vaccination (90%; 64%) at the time of study, which can be explained by the timeline of vaccine rollout in the country that started in mid-February and that faced delays in initiation due to interruptions in receiving vaccine batches into the country.

On the other hand, important results to be highlighted are the severity of illness of COVID-19 associated hospitalization as compared to non-COVID-19 hospitalizations. This study clearly reveals that although COVID-19 hospitalized patients subjectively have a better rating for their general health status, their prognosis is much worse than non-COVID-19 patients hospitalized with longer durations of stay, significantly higher need for intensive care, oxygen therapy and intubation, and ultimately death.

As vaccination may provide false sense of security, our findings highlight the importance of adhering to public health measures to avoid COVID-19 associated hospitalization where vaccinated individuals are still advised to continue practicing hand hygiene, physical distancing, and mask wearing (20).

### Limitations

The study findings are subject to some limitations. First, The MOPH admission database, from which controls were sampled, is not inclusive of the total target population as it covers hospitalization of citizens who are uninsured and who usually belong to the most deprived segments of the population, such as seasonal workers, farmers, retired and unemployed persons, thus on average an older and poorer population (9). Additionally, vaccine effectiveness estimates might be confounded by certain unmeasured behaviors like adherence to non-pharmaceutical interventions, including mask use or recent attendance of gatherings in addition to some variables like socioeconomic status and prior SARS-COV-2 infection. This might have affected our results as uncontrolled confounders might lead to differences in vaccine uptake, exposure to infection, and development of severe disease implications.

## Conclusion

Our study showed that Pfizer-BioNTech vaccine is effective in reducing risk for COVID-19– associated hospitalization in older adults. These findings reinforce the importance of vaccination, among elderly who are at high risk for COVID-19 hospitalization. Additional studies are warranted to explore vaccine effectiveness in reducing hospitalization in younger age groups, as well as reducing COVID-19 infections taking into account other vaccine products in light of the emergence of new SARS-CoV-2 variants and the increase in the elapsed time since vaccination.

## Data Availability

All data produced in the present study are available upon reasonable request to the authors

## Acknowledgments

The authors would like to acknowledge the efforts of the ESU teams in helping in data collection and would like to thank all reporting sites for their continuous collaboration with the MOPH.

## Notes

### Competing Interest Statement

The authors have declared no competing interest.

### Funding Statement

This study did not receive any funding

### Author Declarations

The IRB of the Rafik Hariri University Hospital gave ethical approval for this work

